# What outcome information do women need to support an informed choice between planning a vaginal or caesarean birth? – a consensus study

**DOI:** 10.64898/2026.05.13.26352976

**Authors:** Aniebiet Ekong, Avril Nicoll, Louise Locock, Tara Fairley, Declan Devane, Pauline McDonagh Hull, Lilla Braithwaite, Mo Ade, Ivett Hidvegi, Natalie Saldias, Gillian Taylor, Denitza Williams, Siladitya Bhattacharya, Mairead Black

## Abstract

**Background:** A mode of birth decision aid (DA) can provide information and support discussions about the potential risks and benefits (outcomes) associated with planning a vaginal or caesarean birth. Evidence shows that DAs can enhance patient knowledge, reduce decisional conflict, minimise inconsistencies in decision-making support, especially in maternity settings, and promote informed decision making. Despite these benefits, DAs specific to mode of birth are currently lacking in routine antenatal care. This paper outlines the process we followed to reach consensus on which outcomes of planned mode of birth should be included in a mode of birth DA.

**Methods:** Outcome identification and selection occurred in three phases. Phase 1 involved compiling a long list of outcomes from systematic reviews, the NICE Caesarean Birth Guidance and qualitative interviews with stakeholders. In Phase 2, this list was refined via a 2 round Delphi survey to prioritise outcomes considered important. An outcome reached consensus if ≥70% of all participants, or 70% of women/partners rated it as “critically important” (7–9), and <15% rated it as “not important” (1–3). Phase 3 involved two stakeholder consensus meetings to finalise the outcome list.

**Results:** Seventy-one outcomes were identified. Following two Delphi rounds and consensus meetings, 54 outcomes were rated as critically important. Seventeen outcomes were consistently rated as not critical across both the survey and consensus phases, meaning that ≥70% of participants in each phase did not consider them essential for informing women during pregnancy. Of these, 8 were retained due to NICE recommendations and ultimately, 9 outcomes were excluded. The final list included 44 maternal and 18 child outcomes. Maternal outcomes related to assistance with birth, complications at the time of birth, issues during recovery, pelvic floor, psychological issues, sexual function, and future pregnancy. Child outcomes related to morbidity and death, disease, obesity, issues with cognitive development and physical development.

**Conclusions:** Sixty-two priority outcomes were identified for inclusion in a planned mode of birth DA.

## Introduction

Pregnant women may approach birth with the aim to have either a vaginal birth or a caesarean birth, but the support received to make an informed plan is often suboptimal (1). In the UK, the NICE Caesarean Birth guidance (2,3), and the landmark Supreme Court ruling in *Montgomery vs Lanarkshire* (4) both convey that pregnant women should be provided with information on the potential benefits and risks of planning vaginal or caesarean birth to support informed decision-making (IDM). IDM is considered fundamental to maternity care (5,6). Women who receive comprehensive information regarding their maternity care have reported positive experiences, regardless of the circumstances of their birth (7–9). Furthermore, evidence shows that women can experience improved mental and physical outcomes for themselves and their babies when they are empowered to make informed decisions about their care (10,11). In contrast, a lack of IDM in maternity care has been associated with birth trauma (12). In the UK, information provision is inconsistent, especially in conversations about planned mode of birth (13–15). The UK Birthrights/Mumsnet survey (15) indicated that planned mode of birth “options” and “choices” were discussed haphazardly; 74% of the 1145 women surveyed indicated that they were given the opportunity to discuss the benefits of a vaginal birth, but only 42% were given the opportunity to discuss the benefits of a caesarean birth. One in three women (30%) said that their opinions were not sought in decisions relating to their care. This may reflect longstanding debates about whether women should have the option to choose a caesarean birth and whether or why they want that choice (16). Elements of paternalism may be at play, where the decision to discuss different options with women is based on the assumption that the healthcare provider knows, in absolute terms, what is best for the woman (17). While policy recommendations have moved on, expectations that services should promote vaginal birth may be a persisting factor entrenched in the system, where changing this goes beyond altering individual attitudes and behaviours (18,19).

There is a growing body of literature detailing both the short- and long-term complications of planned and actual mode of birth (20,21) However, it is not clear what information women need in order to make an informed decision between planning vaginal and caesarean birth. The Plan-A project is addressing this issue by developing a decision aid (DA) and implementation guide for use in the UK National Health Service (NHS) to improve routine IDM support for women to plan their mode of birth. A DA outlines the decision to be made, presents the available options, and details the outcomes of each option, including benefits, risks, and uncertainties, while helping people to communicate their values and preferences based on evidence-based information (22). DAs have been shown to increase patient knowledge, reduce decisional conflict, reduce levels of anxiety, lead to IDM, and reduce variability in decision-making support across maternity settings (22–24). The aim of this paper is to explain the processes we undertook to reach a consensus on which outcomes of planned mode of birth to include in the DA.

## Methods

### Outcomes definition

In line with definitions provided by NICE and broader healthcare literature, outcomes have been defined as the measurable effects of an intervention on an individual (25–28). They encompass not only clinical indicators such as diagnoses and complications, but also social, emotional and psychological dimensions that may be influenced by the intervention (25,27). Together, these outcomes contribute to an individual’s overall health-related quality of life. This definition reflects the broad scope of outcomes in healthcare, which includes changes in health status, behaviour and wellbeing. While recognising that evidence on outcomes of planned and actual mode of birth is observational in nature (and not based on randomised trials), this broad definition of outcomes was used for the purpose of this study.

### Outcomes identification

A consensus process was undertaken. The identification of potential outcomes of relevance involved three phases.

1. Phase 1: Development of an initial ‘long-list’ of potentially relevant outcomes from three sources of evidence.
2. Phase 2: A 2-round Delphi survey with stakeholders to update the ‘long-list’, prioritise outcomes, and identify any for which there was consensus on the importance of considering them when planning mode of birth.
3. Phase 3: A consensus meeting with stakeholders to reach an agreement on outcomes of importance to be included in the DA.

### Phase 1- development of an initial ‘long-list’ of potentially relevant outcomes

The development of an initial ‘long-list’ was informed by three sources:

#### Systematic reviews

Potential outcomes were identified from three systematic reviews which are described separately. The first review explored the information requirements, decision support needs and experiences of women planning their MOB. The second review explored the underlying factors influencing women’s birth mode preferences, as well as the key barriers and facilitators to achieving supported decision making in this context. The third review aimed to update the NICE Caesarean Birth guidance on outcomes of planned mode of birth.

#### National Institute for Health and Care Excellence (NICE) caesarean birth guideline

Potential outcomes were identified by extracting outcomes that are recommended for discussion with women during antenatal care in the NICE Caesarean Birth guidance (2). In line with the study protocol, all of these outcomes will be included in the DA. However, these outcomes were also incorporated into the Delphi process to determine whether women considered them critical to be informed of when making their mode of birth plan decision.

#### Qualitative research with stakeholders

Outcomes were also derived from the findings of qualitative interviews conducted with women, partners, and healthcare professionals. Seventy-nine qualitative interviews (including two couples) were conducted in the UK from May 2023 to April 2024, with the wider remit of understanding decision support processes and needs when planning mode of birth. Purposive sampling led to a sample from across the UK of 37 women (≥16 years old) who were currently pregnant, pregnant in the last 10 years, or planning a pregnancy; 5 partners of such women; and 39 NHS health professionals. Participants were recruited using social media platforms, charities, professional organisations, and via email cascades in the five Plan-A NHS study sites. Maximum variation purposive sampling was employed to ensure a diverse representation of the population, capturing a broad range of experiences and characteristics.

Interviews were facilitated by two experienced qualitative researchers (A.E and A.N). Open ended questions explored participants’ experience of mode of birth planning including values and outcomes of importance. Perceived barriers and facilitators to DA use were also explored. Outcomes of importance were extracted and compared with outcomes from the systematic literature reviews. The research team agreed on which outcomes could be combined, and which were new.

### Phase 2- prioritising outcomes of importance and further development of outcomes list using 2-round Delphi surveys

Using standard methodologies from previous studies (29,30), we conducted a two-round modified Delphi online survey and a final consensus meeting to explore and reach consensus on which outcomes are most important to pregnant women. Using multiple rounds of questionnaires, each informed by anonymized feedback from previous rounds, the Delphi methodology is a systematic way to build expert consensus. Through this iterative process participants are encouraged to consider and refine their responses based on the collective input of the group (31–33). The modified Delphi method (which incorporates both surveys and consensus(which incorporates both surveys and consensus meeting) has advantages over the original approach as it offers a more structured communication process, wider participant reach (34), and the opportunity for expert interaction in the final round (35,36).

#### Study population

Participants were recruited from the following stakeholder groups:

1. UK women (≥16 years old) who reported being currently pregnant, pregnant in the last 10 years, or planning a pregnancy
2. Partners of such women
3. NHS health professionals who provide care for pregnant women and their children, including midwives, obstetricians, obstetric anaesthetists and paediatricians. Reflecting the long-term impact of childbirth, we also aimed to recruit from health professional groups such as GPs, health visitors, clinical psychologists, speech and language therapists and pelvic health physiotherapists.

#### Recruitment strategy

To ensure maximal representation, especially from groups whose voices have historically been lacking in research (37), several charities were involved in recruitment. These charities represented reproductive health, pregnancy, childbirth and breastfeeding, and minority ethnic groups. Multiple Maternity and Neonatal Voices partnerships were also involved. Health professionals were identified via networks relevant to maternity care professionals in the UK and via personal contacts. The study advert contained a direct link to the survey and was shared through these organisations and via social media.

Reasons for exclusion from the survey included an inability to consent, and an inability to understand written or spoken English. The latter was a practical step as the survey was conducted in English. Additionally, student midwives, medical students or women/partners who planned birth more than 10 years previously were excluded due to lack of sufficient experience relevant to current birth planning processes.

#### Survey process

Survey questionnaire wording was extensively reviewed internally with our diverse Patient and Public Involvement (PPI) panel to ensure that the eventual wording was accessible and inclusive. Additionally, a think aloud exercise (38) on the questionnaire wording was conducted with 4 participants from diverse backgrounds to garner real time feedback on how potential participants might engage with the survey text, comprehension, and time taken to complete the survey. Think-aloud exercises provide valuable insights into the cognitive processes that occur within working memory in real-time. By collecting various viewpoints and variations in meaning, they have been utilized to understand how research participants from diverse cultural and sociodemographic backgrounds interpret survey questions (39,40). This process helps enhance inclusivity by ensuring that surveys account for a range of interpretations and experiences. The think-aloud process resulted in changes to the wording and an indication of time taken to complete the survey.

Survey participants were presented with sociodemographic questions and a list of outcomes with their descriptions arranged in two domains: maternal outcomes and child outcomes. The wording for descriptions was determined by the research team (Chief investigator, research fellows and PPI lead) and members of the PPI panel. The aim was to make the language used for descriptions as simple as possible to foster inclusivity.

The Delphi survey was conducted using REDCap electronic data capture software hosted on the University of Aberdeen server (41,42). REDCap (Research Electronic Data Capture) is a secure, web-based platform designed to facilitate data collection for research studies (41). The first round of the Delphi survey was opened from 11^th^ January 2024 to 25^th^ of February 2024. A list of 55 outcomes were presented in round 1 of the survey, and participants were invited to rate each outcome on a 9-point, 3 domain Likert scale, where 9 indicated “critical” as being the highest level of importance for discussion ahead of planning mode of birth and 1 indicated “not important”. The 9-point Likert scale has been deemed as advantageous over the 5- or 3-point Likert scales in preliminary stages of Delphi studies that prioritize outcomes as it leads to a broader set of outcomes being included (43).

The 3 domains categorized the Likert points into “critical” (points 7-9), “important but not critical” (points 4-6) and “not important” (points 1-3). Participants were invited to suggest additional outcomes. New suggestions relevant to mode of birth planning were subsequently included in round 2 for rating. Following closure of round 1, frequency distribution and median ranges of ratings for each outcome by each stakeholder group was calculated using the IBM SPSS Statistics software (Version 29).

The second survey was opened from 19^th^ April 2024 to 16^th^ May 2024. Participants who took part in round 1 were sent personalised invitation emails to take part in the second round. No new participants took part in round 2. Participants were then presented with a percentage distribution of scores for each point on the 9-point scale, according to stakeholder groups in the previous round, along with their previous rating for the outcome. Participants were asked to review their rating from round 1, including considering whether they intended to change their initial scores for each outcome. The same Likert scale was used in round 2. All original outcomes from round 1 were presented in round 2, with an additional 21 outcomes from round 1 suggestions and the updated analysis of qualitative interview transcripts. As predefined in the protocol, an outcome was considered to have reached consensus if ≥ 70% of all participants or, failing that, ≥ 70% of women/partner participants rated it as “*critically important*” (7–9), and fewer than 15% rated it as “*not important*” (1–3).

### Phase 3- consensus meeting to reach an agreement

The final stage of the planned mode of birth outcomes development process was a virtual open forum consensus meeting over 2 half days. Women, partners and health professionals who met the study population criteria (2.2.1) and had previously indicated interest were invited to participate as the ‘expert panel’ in the consensus meetings. Nineteen stakeholders consisting of 5 members of the research team, 1 PPI partner, 5 women, and 8 health professionals attended day 1 of the meeting, and 16 stakeholders attended day 2: 4 members of the research team, 1 PPI partner, 5 women, and 6 health professionals. Two health professionals and two members of the research team who attended day 1 of the consensus meeting were absent on day 2. However, one additional member of the research team joined on day 2. Of all the stakeholders attending, only the study participants were eligible to vote in the consensus meeting. To address potential power imbalances and the risk that women might feel discouraged from speaking openly in the presence of health professionals, skilled facilitators were used to guide the discussions. Additionally, recognising that modified Delphi method can reduce anonymity, anonymous voting was incorporated to preserve participants privacy.

Forty-nine outcomes that received critical support (i.e., were rated as critical by participants) in the survey rounds were presented one-by-one to stakeholders for discussion. This allowed them to review the ratings, assess whether they agreed with them, and suggest potential outcome combinations. Additionally, 22 outcomes that did not receive critical support were presented for discussion, in turn, during which study participants re-evaluated them. Of these 22 outcomes, 4 were ultimately rated as critical for inclusion in the DA. Three outcomes, changes in sexual function following childbirth, duration of labour, and cephalopelvic disproportion, were included based solely on votes from recruited women, while one outcome, scratches and bruises on the baby’s head, was included based on votes from all study participants. A decision was made to repoll the rating of a fifth outcome, caesarean scar ectopic pregnancy, which had received 69.2% of votes as critical on the first day of the meeting. On the second day, a decision was reached to include it in the DA. as part of a combined outcome with placenta accreta. All other outcomes that were not rated as critical in the surveys remained non-critical in the consensus meeting. Figure 1 (supplementary material) shows a flowchart of the Delphi process to identify outcomes for inclusion in the Plan-A DA.

**Figure 1:**
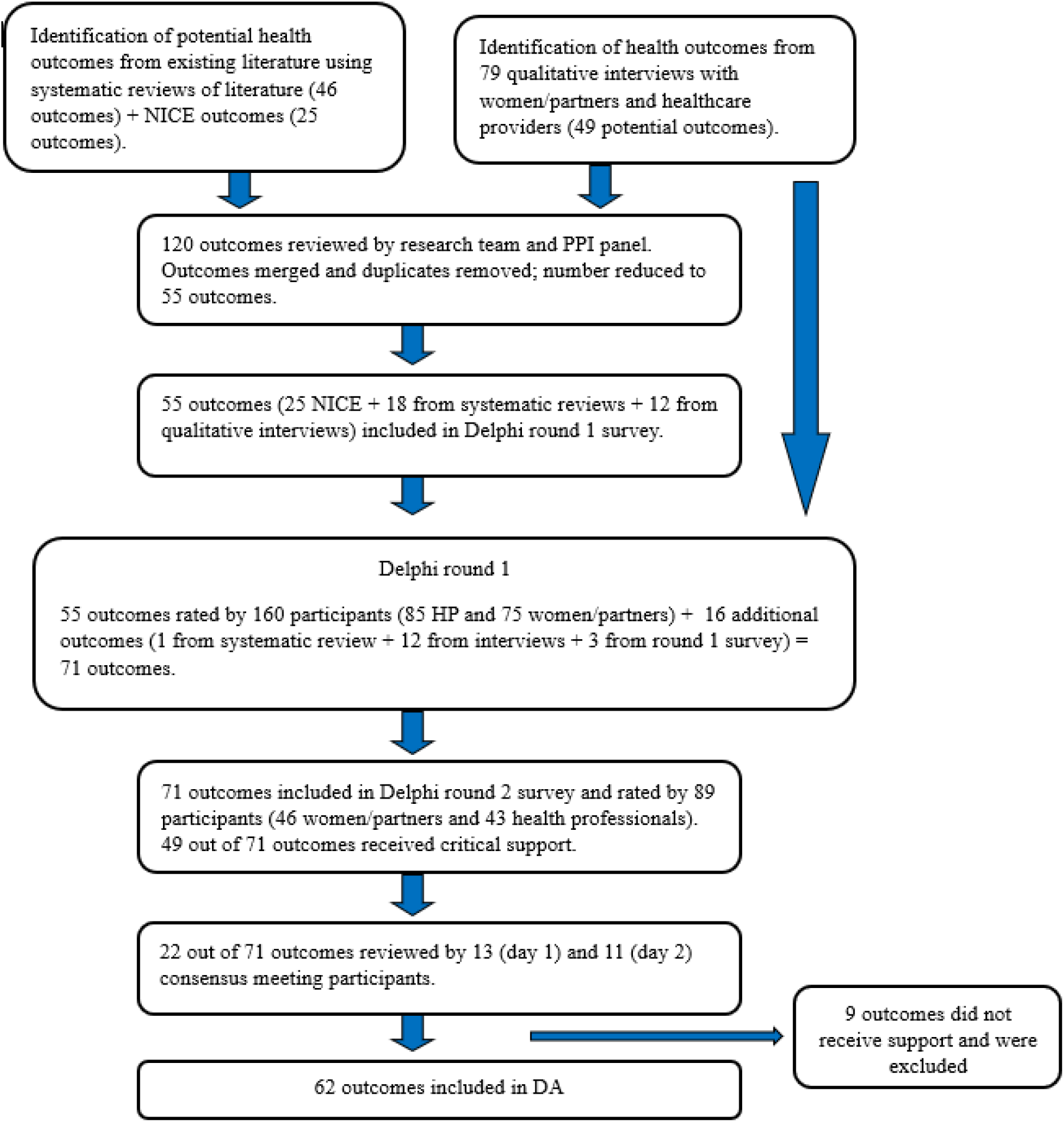
Flowchart of the Delphi process to identify outcomes for inclusion in the Plan-A DA.

#### Ethics approval

Ethics approval for this research was provided by the East of Scotland Research Ethics Service and the Health Research Authority (HRA) and Health and Care Research Wales (HCRW).

## Results

### Phase 1 - developing an initial ‘long-list’ covering potentially relevant outcomes

#### A.) Systematic reviews

Thirty-three studies reporting data for at least 2764 participants in Europe, North America (USA and Canada), Australia, Taiwan and Japan, were included in the review exploring the information requirements and decision support needs of women planning MOB. The second review that explored factors influencing women’s birth mode preferences reported data in 46 studies for at least 4663 participants in Europe (including the UK), North America (USA and Canada), Australia and Japan.

Forty-six outcomes of importance were identified from three systematic reviews and assessed for inclusion in the Delphi surveys (phase 2). Following a process of merging and relabelling, 19 outcomes relevant to planned mode of birth were selected for inclusion across both survey rounds (18 in survey round 1, and 1 added in survey round 2; see Table 1).

**Table 1:** Outcomes identified and included from different sources.

| **NICE outcomes** | **Systematic reviews** | **Qualitative interviews** | **Survey round 1** |
| --- | --- | --- | --- |
| Peripartum hysterectomy | Chances of a vaginal birth | Pelvic organ prolapse | Childhood cancer risk |
| Maternal death | Scar healing/physical appearance | Immediate skin to skin contact | Coccyx pain |
| Length of hospital stay | Duration of labour | Delayed cord clamping | Coeliac disease |
| Placenta accreta in a future pregnancy | Receiving advice on the impact of medical conditions on childbirth | Back pain |  |
| Uterine rupture in a future pregnancy | Cephalo-pelvic disproportion | Placenta praevia in a future pregnancy |  |
| Urinary incontinence | The chance of a vaginal birth in a future pregnancy after a caesarean birth | Psychosis following birth |  |
| Faecal incontinence | Postpartum recovery | Post traumatic stress disorder due to childbirth |  |
| Vaginal tear | Higher surgical risk with each caesarean section | Ventouse birth |  |
| Pain during childbirth | The chance of a scar rupture relative to the time since previous birth | Analgesia/anaesthesia complications |  |
| Thromboembolic disease | Recommended pregnancy spacing after a previous caesarean section | Future fertility |  |
| Major obstetric haemorrhage | Caesarean process | Postpartum anxiety |  |
| Postnatal depression | Forceps birth | Breastfeeding initiation |  |
| Admission to neonatal unit | Placental abruption | Risk of preterm birth in a future pregnancy |  |
| Infection in babies | Recovery after episiotomy | Vaginal birthing process |  |
| Persistent verbal delay | Shoulder dystocia | Caesarean scar ectopic pregnancy |  |
| Infant mortality | Changes in sexual function following childbirth | Meconium aspiration syndrome |  |
| Neonatal mortality | Chance of having an emergency caesarean section | Hypoxic ischaemic encephalopathy |  |
| Asthma | Episiotomy | Scratches and bruises on baby's head |  |
| Childhood obesity | Stillbirth risk | Short- or long-term nerve damage |  |
| ITU admission |  | Emotions experienced in relation to birth |  |
| Stillbirth in a subsequent pregnancy |  | Ileus |  |
| Respiratory morbidity |  | Satisfaction with birth experience |  |
| Cerebral palsy |  | Infection (mother) |  |
| Autism spectrum condition |  | Satisfaction with birth experience |  |
| Type 1 diabetes |  | Sensory cognitive development |  |

Twenty-five outcomes were included from the 2021 NICE Caesarean Birth guidance on the benefits and risks of caesarean and vaginal birth (See table 1).

#### B.) Qualitative interviews/outcomes

##### Women and partners

The interview sample of 42 individuals comprised 76% White participants, 12% Asian, 10% African, and 2% of mixed ethnicity. Participants were recruited from England (60%), Scotland (21%), Wales (2%) and Northern Ireland (2%), while location data was unreported for 14% of the sample. Most of the participants were between the ages of 35 and 39 years at interview (40%), while 7% were under the age of 25. The demographic characteristics of the sample, including diversity of age, sexual orientation, relationship status, religion and household income are detailed in Appendix Table 1.

##### Health professionals

A total of 39 health professionals were interviewed, made up of obstetricians (11), midwives (16), anaesthetists (3), GPs (3), paediatricians (2), a clinical psychologist (1), a family nurse (1), a pelvic health physiotherapist (1), and a health visitor (1).

##### Outcomes from interviews

Sixty-one potential outcomes were identified from interviews with women/partners and health professionals. After removal of duplicates and merging with outcomes from ongoing systematic reviews and interviews, 24 outcomes were considered for inclusion in both rounds of the survey. Table 1 includes the list of outcomes included in the survey.

### Phase 2- Delphi survey process and outcomes

Table 2 shows the characteristics of all survey participants. In the first survey, 160 participants took part: 70 women, 5 partners, and 85 health professionals. The majority of participants were from Scotland (74) and England (72). There were 9 participants from Wales and 5 from Northern Ireland. One hundred and twenty-four participants in the survey were ≥ 35 years, 34 participants were age 25-34 years, and 2 participants were age less than 25 years. Among the women/partner participants, 51 participants identified as White British, 14 as White other, 4 as Black African and 4 as Asian.

**Table 2:** Characteristics of all survey participants (women/partners and health professionals)

| **Characteristics** | **Survey 1** | **Survey 2** |
| --- | --- | --- |
| ***Age range*** |  |  |
| Less than 25 years | 2 | 2 |
| 25-34 years | 34 | 20 |
| 35-39 years | 57 | 27 |
| 40 years and older | 67 | 40 |
| ***Sexual orientation*** |  |  |
| Bisexual | 10 | 5 |
| Gay or lesbian | 3 | 3 |
| Straight/heterosexual | 144 | 80 |
| Prefer not to say | 3 | 1 |
| ***Relationship status*** |  |  |
| Civil partnership | 5 | 2 |
| Partnered | 34 | 18 |
| Divorced | 4 | 2 |
| Married | 111 | 64 |
| Single | 6 | 3 |
| ***Ethnicity*** |  |  |
| Asian/Asian British | 5 | 3 |
| Black | 7 | 2 |
| Mixed | 2 | 2 |
| White British | 119 | 64 |
| White other | 24 | 16 |
| Other | 3 | 2 |
| ***Religion*** |  |  |
| Agnostic | 9 | 6 |
| Christianity | 63 | 36 |
| Islam | 5 | 3 |
| Hinduism | 1 | - |
| None | 76 | 40 |
| Other religion | 4 | 3 |
| Prefer not to answer | 2 | 1 |
| ***Education*** |  |  |
| Foundation qualifications | 6 | 4 |
| Advanced qualification | 4 | 4 |
| Higher qualification | 146 | 80 |
| Prefer not to answer | 5 | 1 |
| ***Income status*** |  |  |
| Less than £20,000 | 4 | 3 |
| £20,000 -£39,000 | 14 | 7 |
| £40,000-£59,000 | 29 | 15 |
| More than £60,000 | 105 | 61 |
| Prefer not to answer | 5 | 1 |
| ***Living status*** |  |  |
| Asylum seeker | 1 | - |
| Prefer not to answer | 1 | - |
| ***Country of residence*** |  |  |
| Scotland | 74 | 46 |
| Wales | 9 | 3 |
| England | 72 | 38 |
| Northern Ireland | 5 | 2 |
| ***Previous birth experiences and current status*** | ***Survey 1*** | ***Survey 2*** |
| Planned caesarean section | 16 | 13 |
| Unplanned/emergency C.S | 15 | 11 |
| Spontaneous vaginal birth | 18 | 14 |
| Assisted vaginal birth (forceps, suction cup, ventouse) | 13 | 13 |
| Currently pregnant | 11 | 10 |

Eighty-nine participants from 160 that took part in survey round 1 completed survey round 2: 44 women, 2 partners, and 43 health professionals; with 46 participants from Scotland, 38 from England, 3 from Wales, and 2 from Northern Ireland.

The overall attrition rate was 44%; 38% for women/partners and 49% for health professionals.

#### Outcomes

Fifty-five outcomes categorized into maternal outcomes and child outcomes were included in round 1 for rating; 38 out of those 55 outcomes received critical support. In round 2, 71 outcomes were included: 55 outcomes from survey round 1 and 16 additional outcomes from ongoing interviews (12 outcomes), systematic reviews (1 outcome), and survey round 1 (3 outcomes. After round 2, 49 out of 71 outcomes received critical support. A list of all 71 outcomes included in both survey rounds can be found in Table 2 (appendix). All outcomes were presented in the Delphi study as listed in Table 2 (appendix) with their explanations, without indicating whether they were more, less, or equally likely to occur with planned caesarean birth or planned vaginal birth.

### Phase 3- Consensus meeting

Among the five women who participated in the consensus meeting, one was currently pregnant, and all others had previously given birth. Among those who had previously given birth, one woman had an assisted vaginal birth, two had a planned caesarean birth and one had an unplanned caesarean birth. The women’s ages ranged from 25 years to over 40 years old. In addition, all the women were educated to a higher qualification (Bachelors, Masters, higher diploma or equivalent). The health professionals included 3 midwives and 6 consultant obstetricians, 1 of whom specialized in foetal maternal medicine.

The criteria for outcome inclusion were predefined in the study protocol (stated in 2.2.4) and applied in both survey rounds and the consensus meeting. To address the numerical imbalance between health professionals and women participants, outcomes were also included if at least 4 out of 5 (80%) of the women participants rated them as critical. All outcomes that received critical support remained consistent across survey rounds. Participants at the consensus meeting selected and prioritised 4 of the remaining 22 outcomes to be included in the DA. Additionally, one outcome was repolled due to being on the border of achieving critical support, leading to an agreement to include it as a combined outcome with another.

While ratings for several outcomes were similar between health professionals and women/partners, differences emerged for some outcomes. “Changes in sexual function following childbirth”, “duration of labour”, and “cephalopelvic disproportion” were rated as critical by women/partners but were not considered critical by health professionals. Conversely, “caesarean scar ectopic pregnancy” was prioritized as critical by health professionals but not by women/partners. Health professionals raised the possibility that women may not have had a full understanding of the gravity of the outcome and whether that might have influenced women’s voting in the survey. However, the efforts made to inform women about the meaning of each outcome during the survey rounds and ahead of consensus meetings were noted and felt, as a group, to be adequate to convey their meaning. Additionally, survey responses indicated that health professionals regarded “scratches and bruises on the baby’s face” as a critical outcome for inclusion, while women did not, prompting health professionals to question whether routine discussion of this outcome should continue in current clinical practice.

In line with the study protocol, eight outcomes recommended by NICE for discussion with women in the NICE Caesarean Birth guidance were included in the DA, even though they were not rated as critical by the study participants.

In general, the consensus meeting discussions largely focused on the potential combination of outcomes and suggestions for wording of certain outcomes. For instance, participants suggested the inclusion of other significant types of vaginal trauma other than 3^rd^ and 4^th^ degree tears that, while not involving the sphincter muscles, can be debilitating and challenging for women. This led to a broadening of the definition of vaginal tears to include both third and fourth-degree tears and these additional injuries. There were also suggestions for the potential combination of placenta accreta with placenta previa and caesarean scar ectopic pregnancy, and episiotomy with recovery from episiotomy. Consensus was reached to change from ‘delayed’ cord clamping to ‘deferred’ cord clamping to align with RCOG’s current terminology (44).

Seventeen outcomes were rated as not critical in the survey rounds and the consensus meeting. This means that 70% of participants in both phases did not consider these outcomes essential for women to be informed about during pregnancy. However, in line with the study protocol, 8 of these 17 outcomes were retained due to being recommended by NICE for discussion ahead of birth. Ultimately, 9 outcomes were considered not critical to include in the DA. The excluded outcomes were scar healing/physical appearance, the chance of a vaginal birth in a future pregnancy after a caesarean birth, ileus, coccyx pain, back pain, psychosis following birth, coeliac disease, childhood cancer risk and sensory cognitive development.

Table 3 outlines the list of 22 outcomes discussed during the consensus meeting and decisions taken and Table 4 outlines the 62 outcomes included in the DA.

**Table 3:** List of 22 outcomes discussed during the consensus meeting and decisions reached.

| **Outcomes** | **Rating** | **Comments/final decision** |
| --- | --- | --- |
| Length of hospital stay | Exclude | Include as NICE recommends |
| Pain during/after childbirth | Exclude | Include as NICE recommends |
| The chance of a vaginal birth in a future pregnancy after a caesarean birth. | Exclude | Exclude |
| Caesarean scar ectopic pregnancy | Missed the critical mark (69.2%) | Include as combination with placenta accreta |
| Ileus | Exclude | Exclude |
| Changes in sexual function following childbirth | Critical | Include |
| Duration of labour | Critical | Include |
| Cephalopelvic disproportion | Critical | Include |
| Intensive care admission | Exclude | Include as NICE recommends |
| Scar healing/physical appearance | Exclude | Exclude |
| Coccyx pain | Exclude | Exclude |
| Persistent verbal delay | Exclude | Include as NICE recommends |
| Childhood asthma | Exclude | Include as NICE recommends |
| Childhood obesity | Exclude | Include as NICE recommends |
| Autism spectrum condition | Exclude | Include as NICE recommends |
| Back pain | Exclude | Exclude |
| Psychosis following birth | Exclude | Exclude |
| Childhood cancer risk | Exclude | Exclude |
| Coeliac disease | Exclude | Exclude |
| Sensory cognitive development | Exclude | Exclude |
| Type 1 diabetes | Exclude | Include as NICE recommends |
| Scratches and bruises on baby's head | Critical | Include |

**Table 4:**
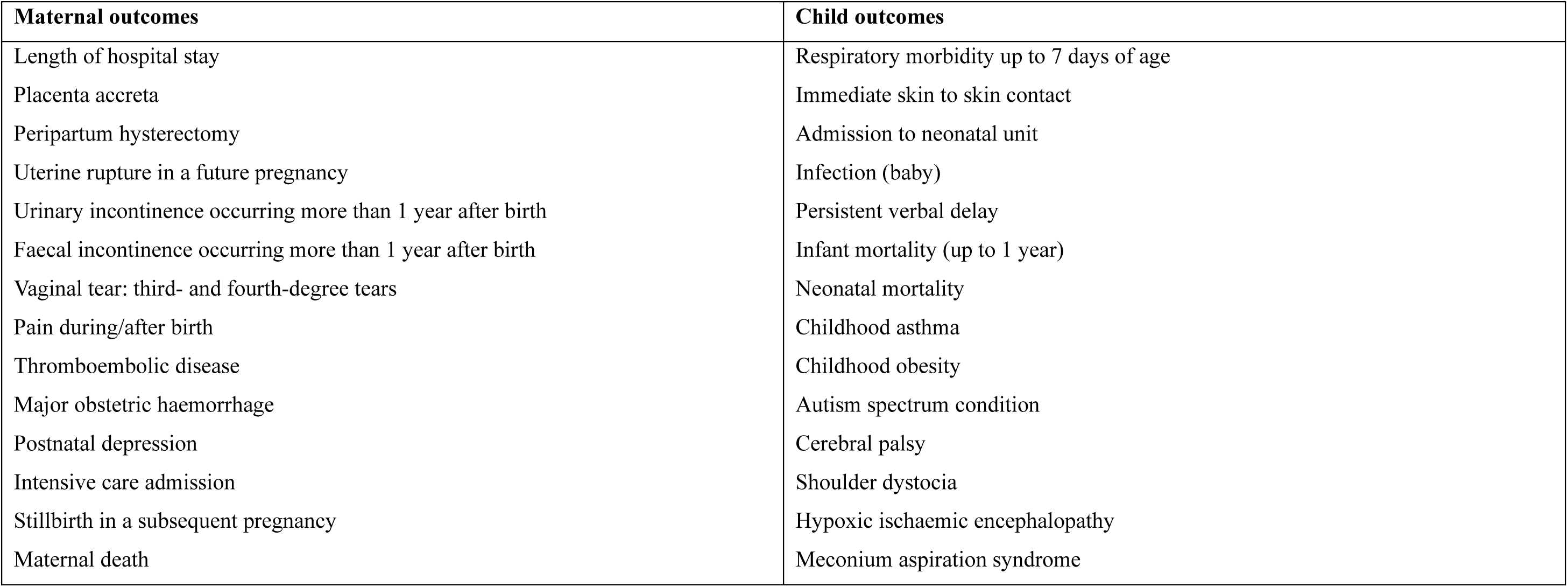
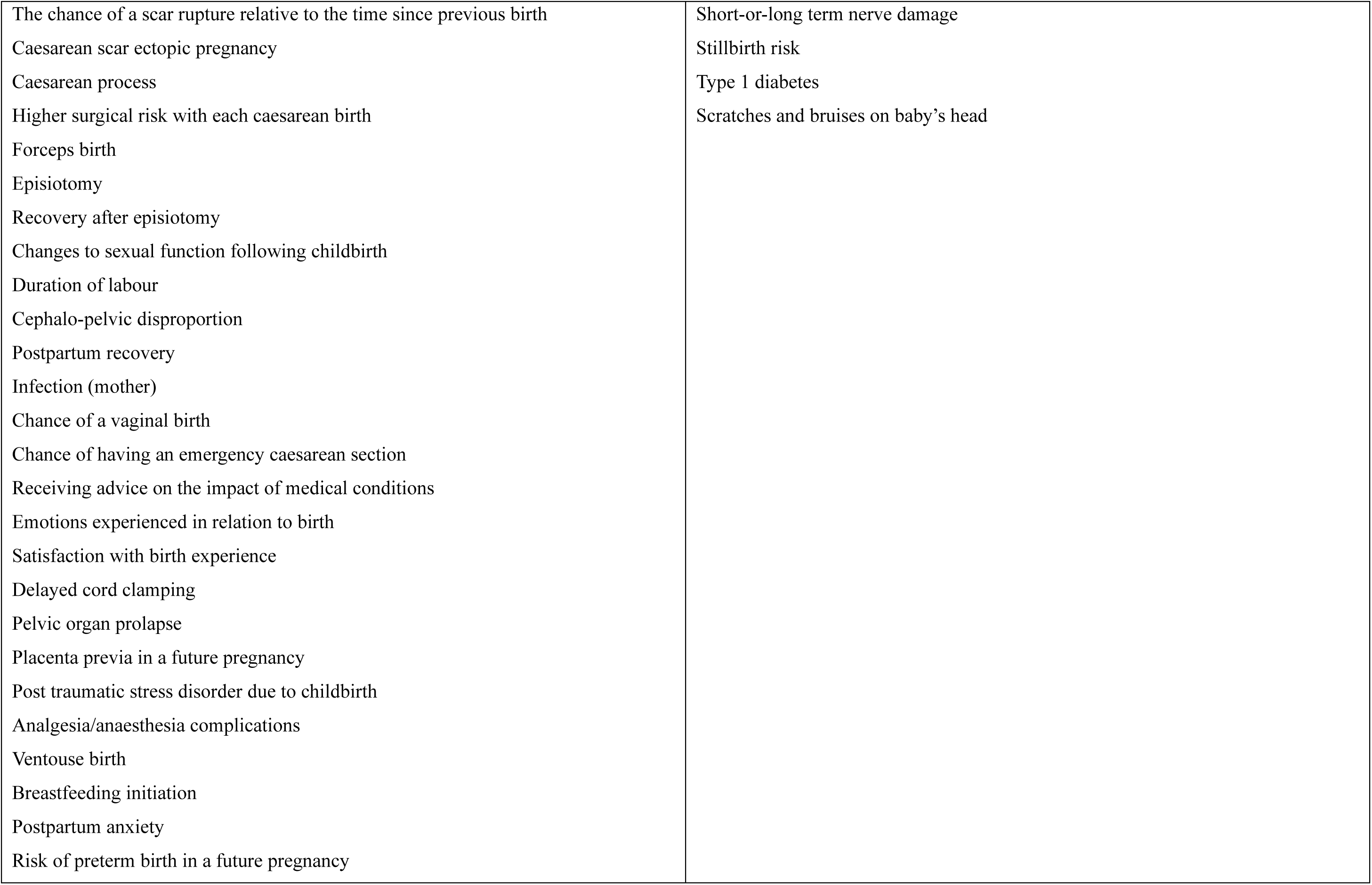
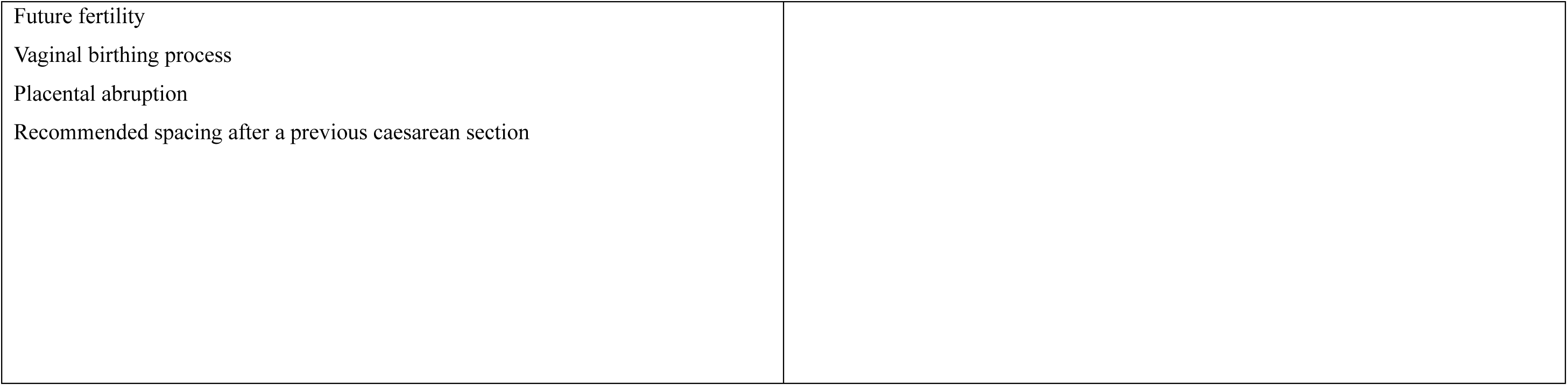
List of outcomes included in the DA.

| **Maternal outcomes** | **Child outcomes** |
| --- | --- |
| Length of hospital stay | Respiratory morbidity up to 7 days of age |
| Placenta accreta | Immediate skin to skin contact |
| Peripartum hysterectomy | Admission to neonatal unit |
| Uterine rupture in a future pregnancy | Infection (baby) |
| Urinary incontinence occurring more than 1 year after birth | Persistent verbal delay |
| Faecal incontinence occurring more than 1 year after birth | Infant mortality (up to 1 year) |
| Vaginal tear: third- and fourth-degree tears | Neonatal mortality |
| Pain during/after birth | Childhood asthma |
| Thromboembolic disease | Childhood obesity |
| Major obstetric haemorrhage | Autism spectrum condition |
| Postnatal depression | Cerebral palsy |
| Intensive care admission | Shoulder dystocia |
| Stillbirth in a subsequent pregnancy | Hypoxic ischaemic encephalopathy |
| Maternal death | Meconium aspiration syndrome |
| The chance of a scar rupture relative to the time since previous birth<br>Caesarean scar ectopic pregnancy<br>Caesarean process<br>Higher surgical risk with each caesarean birth<br>Forceps birth<br>Episiotomy<br>Recovery after episiotomy<br>Changes to sexual function following childbirth<br>Duration of labour<br>Cephalo-pelvic disproportion<br>Postpartum recovery<br>Infection (mother)<br>Chance of a vaginal birth<br>Chance of having an emergency caesarean section<br>Receiving advice on the impact of medical conditions<br>Emotions experienced in relation to birth<br>Satisfaction with birth experience<br>Delayed cord clamping<br>Pelvic organ prolapse<br>Placenta previa in a future pregnancy<br>Post traumatic stress disorder due to childbirth<br>Analgesia/anaesthesia complications<br>Ventouse birth<br>Breastfeeding initiation<br>Postpartum anxiety<br>Risk of preterm birth in a future pregnancy | Short-or-long term nerve damage<br>Stillbirth risk<br>Type 1 diabetes<br>Scratches and bruises on baby's head |
| Future fertility<br>Vaginal birthing process<br>Placental abruption<br>Recommended spacing after a previous caesarean section |  |

#### PPI involvement

The study’s PPI partners played a vital role in shaping the study. They advised on the language used in study materials, including the survey questionnaire, participant information sheets and consent forms, to ensure these were accessible to a lay audience. They also supported community engagement efforts, which helped to build trust and contributed significantly to the recruitment process.

PPI partners provided valuable insight into the qualitative data from women and health professionals, challenged our initial interpretations and helped to contextualize the findings, which strengthened the overall analysis.

## Discussion

The main objective of this paper was to outline the process of achieving consensus on what information on outcomes of planned mode of birth to include in a DA. These outcomes represent the consensus opinion of 70 women, 5 partners and 85 healthcare professionals across the UK. Consensus was reached to include 54 out of 71 outcomes, with an additional 8 outcomes included based on the NICE Caesarean Birth guidance recommendations (2). The final outcomes list included 62 outcomes, comprising both maternal (44) and child (18) outcomes. Maternal outcomes related to assistance with birth, complications at the time of birth, issues during recovery, future pregnancy, pelvic floor, psychological issues, and sexual function. Child outcomes related to morbidity and death, disease, obesity, issues with cognitive development and physical development.

Although a wide range of outcomes related to planned mode of birth have been reported in published literature (20,45), this study identified additional outcomes that may hold particular relevance to women when making mode of birth decisions, outcomes that may be less frequently prioritised by researchers rather than to researchers.

In addition to the outcomes recommended by NICE for planned caesarean birth (2), this consensus process identified 37 outcomes considered important by women, their partners and health professionals when planning mode of birth. Notably, eight outcomes prioritised by NICE for discussion during antenatal care were not rated as critical by participants in this study (Table 3). These findings highlight a broader range of informational needs than those currently reflected in the NICE Caesarean Birth guideline, suggesting that women’s preferences during birth planning may extend beyond existing guidance (2). This information has been systematically compiled using a transparent, documented development process, involving a diverse group of stakeholders, including PPI partners. It is important to acknowledge that there is some uncertainty as to whether outcomes identified as critical would have retained this rating if participants had been informed whether differences existed between both planned modes of birth and the outcomes in question.

A comparable Delphi methodology was utilised in a French study (46) to inform the development of a shared decision-making tool that captures women’s concerns related to childbirth. Consistent with the findings of the present study, key areas of concern identified included the vaginal birth process, strategies for pain relief, and perineal tears. The inclusion of women in both studies, along with the thematic focus on issues of importance to them, underscores a growing demand for IDM in matters concerning their health and maternity care.

A key strength of this study lies in its adherence to a systematic development process for decision aids, aligned with the International Patient Decision Aid Standards (IPDAS) Collaboration criteria for the content specification phase (47–49), as well as the NICE standards framework for shared decision-making support tools (50). Furthermore, the use of a modified Delphi approach enabled a comprehensive exploration of the decisional needs of diverse stakeholders, incorporating perspectives from both women and healthcare professionals and allowed an opportunity for an open forum discussion between women and health professionals. This allowed members of the panel to provide further clarification on some matters and present arguments to justify their viewpoints.

However, although each bring differing but valid perspectives, open forum discussions between health professionals and women can potentially introduce power imbalances that may inhibit open and equitable dialogue (51). In this study, such imbalances may have been further compounded by unequal representation of stakeholder groups at the consensus meeting, where five women/partners and eight health professionals were in attendance. To minimise the influence of such dynamics, several mitigation strategies were implemented. This included involving a public panel member to amplify public voices if they felt this was necessary, appointing a chair and facilitator who were experienced in leading multi-stakeholder meetings, allocating structured time to ensure the women could express their views on each outcome without interruption or influence and promoting anonymous voting. Although the voting system had been tested beforehand, technological challenges meant that a number of participants opted to waive their anonymity when voting. As such, it is acknowledged that some degree of power dynamics may still have influenced how individuals contributed to the discussions or cast their votes.

The development process of this DA prioritized a broad representation of diverse stakeholders, particularly from marginalized communities. In the interview study, 24% of women/partner participants were from ethnic minority backgrounds, and 29% had a household income of less than £20,000. Similarly, in the survey, 13.3% of women/partner participants were from ethnic minority backgrounds, and 5.3% had a household income below £20,000. These figures highlight the inclusion of diverse perspectives in understanding decision support needs. The final open forum consensus meeting involved a less diverse sample, raising the possibility that outcomes particularly important to women from lower socioeconomic and ethnic minority backgrounds may have been underrepresented or excluded. Additionally, the high attrition rate between the two surveys might mean that we were unable to represent more diverse views of mothers. Attrition in general has been identified as a major issue in Delphi studies due partly to the time difference between the launch of the first survey and subsequent rounds and participants’ other time commitments (52,53). However, given that the consensus process was cumulative, beginning with the initial survey and culminating in the final consensus meeting, it is assumed that earlier stages of the process provided opportunities for the inclusion and representation of perspectives from these groups. The study also involved a diverse group of PPI panel members who contributed to setting priorities and research questions, study design, recruitment strategies, and the use of plain and accessible language.

In spite of the diversity of our sample and the systematic process we followed, it is possible that another sample may have come to some different conclusions about what outcomes are critical to include. Some members of our team, for example, were surprised that scar healing/appearance, chance of vaginal birth after caesarean and coccyx/back pain did not reach the threshold for inclusion. For practical reasons, not every possible outcome can be included, and other outcomes may still be important to individual women. This highlights the importance of the DA being a ‘living’ tool, which can evolve through testing in clinical practice, regular review with stakeholders, and as new, higher quality evidence about outcomes of planned mode of birth emerges.

### Strengths and limitations

The use of a modified Delphi method represents a vital strength of this study. This approach promotes anonymity among participants during the questionnaire rounds, thereby reducing the risk of individual dominance and allowing participants to reconsider and refine their responses over successive rounds. It also facilitated the development of a comprehensive list of outcomes, informed by existing literature and stakeholder interviews, and reflective of current perspectives on outcomes important to women. The expert panel included women, their partners, and healthcare professionals, bringing together diverse viewpoints shaped by varying experiences and professional backgrounds.

However, the study is not without limitations. In addition to the issues of power imbalance described above, the response rate declined to 56% in the second survey round, and only 13 experts attended the final consensus meeting. This minimises the continuity between stages of the work, limiting how much of the intended Delphi methodology could be applied across stages, and also limits the range of representation within the consensus group. However, within the limits of the group number, diversity in the sample was still evident,.

Additionally, as outlined in Section 2.2.4, outcomes were presented with descriptive help text only, without comparative data. This approach was taken because the underlying evidence is constantly evolving, sometimes contentious, and may vary depending on the populations being compared. Including comparative data risked oversimplifying or misrepresenting complex evidence. However, this absence of contextual data may have influenced how participants prioritised outcomes, particularly those that may differ only marginally between birth modes.

## Conclusion

We have developed and described the process used to achieve a consensus on mode of birth outcomes relevant to women, their partners and health professionals involved in their care to include in a DA for planning mode of birth in routine maternity care. Other aspects of developing the DA will be described in future papers.

## Data Availability

All data generated or analysed during this study are included in this published article and the supplementary information files available from the OSF repository.

https://osf.io/5d69b/overview

## List of abbreviations

DA: Decision Aid
HCRW: Health and Care Research Wales
HRA: Health Research Authority
IBM: International Business Machines
IDM: Informed Decision Making
IPDAS: International Patient Decision Aid Standards
MOB: Mode of Birth
NICE: National Institute for Health and Care Excellence
NHS: National Health Service
PPI: Patient and Public Involvement
REDCAP: Research Electronic Data Capture
SPSS: Statistical Package for the Social Sciences
UK: United Kingdom

## Statements and Declarations

### Ethics approval and consent to participate

Informed consent was obtained from independent study participants prior to their involvement in the study.

### Consent for publication

Written informed consent was obtained for the publication of anonymised patient information included in this article

### Competing interests

Pauline Hull is the founder of the Caesarean Birth Charity and a co-author of *Choosing Cesarean: A Natural Birth Plan* (Prometheus Books, 2012), and Mairead Black was a member of the working group developing decision aids as part of the NHS England Maternity Transformation Programme Personalisation and Choice stream (2021-2024); neither considers these roles to be competing interests, and neither influenced the results or discussion reported in this manuscript

### Funding

This study was funded by the UK National Institute for Health and Care Research (NIHR) Health and Social Care Delivery Research (HSDR) Programme (award ID: NIHR150979), following external peer review by scientific, clinical, and public experts. The funder had no role in any part of the research process. The funder had no role in the design, conduct, analysis, or reporting of the study, including the decision to submit for publication.

### Author contributions

MB conceived the idea for the project, contributed to the development of the protocol, study design and conduct, and revised manuscript drafts. AE developed study materials, designed and conducted the survey, collected and analysed data and wrote the first and subsequent drafts of the manuscripts. AN contributed to study design, collected and analysed qualitative data and revised manuscript drafts. LL,TF, PMH contributed to the study concept and study design, development of the protocol, interpretation of the qualitative data and editing of the manuscript. DD contributed to the development of the protocol, survey study design and contributed to revisions of the manuscript. MA, LB, IH, NS, GT and DW contributed to the design of study documents and interpretation of study findings. SB contributed to the study design and was one of the facilitators at the consensus meeting. All authors had an opportunity to edit and approve the manuscript.

## Acknowledgements

We would like to thank VE and PMH for their support in reviewing and refining the manuscript, and for their contributions as members of the research team. We are also deeply grateful to all the Patient and Public Involvement (PPI) contributors to the Plan-A study, as well as to all participants in the interview and consensus studies, for their generous time and input.

We acknowledge the charity AIMS for their valuable involvement in the interpretation of study’s findings. Special thanks to Gillian Ferry, the study’s coordinator for her valuable support.

## Creative Commons Attribution

For the purpose of open access, the author has applied a Creative Commons Attribution (CC BY) licence to any Author Accepted Manuscript version arising from this submission.

## Appendix

**Table S1:**
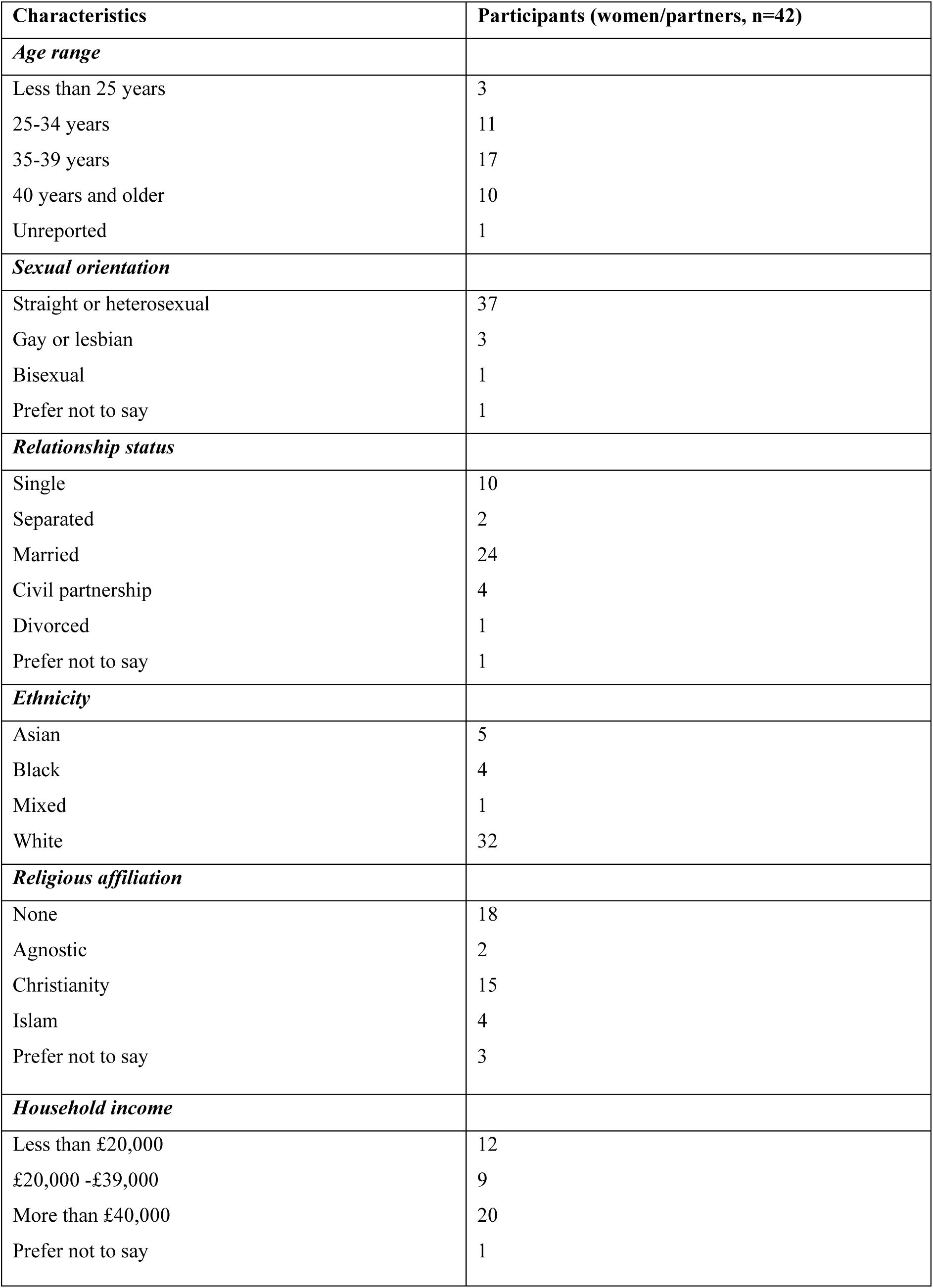
Characteristics of women/partner interview participants.

| Characteristics | Participants (women/partners, n=42) |
| --- | --- |
| ***Age range*** |  |
| Less than 25 years | 3 |
| 25-34 years | 11 |
| 35-39 years | 17 |
| 40 years and older | 10 |
| Unreported | 1 |
| ***Sexual orientation*** |  |
| Straight or heterosexual | 37 |
| Gay or lesbian | 3 |
| Bisexual | 1 |
| Prefer not to say | 1 |
| ***Relationship status*** |  |
| Single | 10 |
| Separated | 2 |
| Married | 24 |
| Civil partnership | 4 |
| Divorced | 1 |
| Prefer not to say | 1 |
| ***Ethnicity*** |  |
| Asian | 5 |
| Black | 4 |
| Mixed | 1 |
| White | 32 |
| ***Religious affiliation*** |  |
| None | 18 |
| Agnostic | 2 |
| Christianity | 15 |
| Islam | 4 |
| Prefer not to say | 3 |
| ***Household income*** |  |
| Less than £20,000 | 12 |
| £20,000 -£39,000 | 9 |
| More than £40,000 | 20 |
| Prefer not to say | 1 |

**Table S2:**
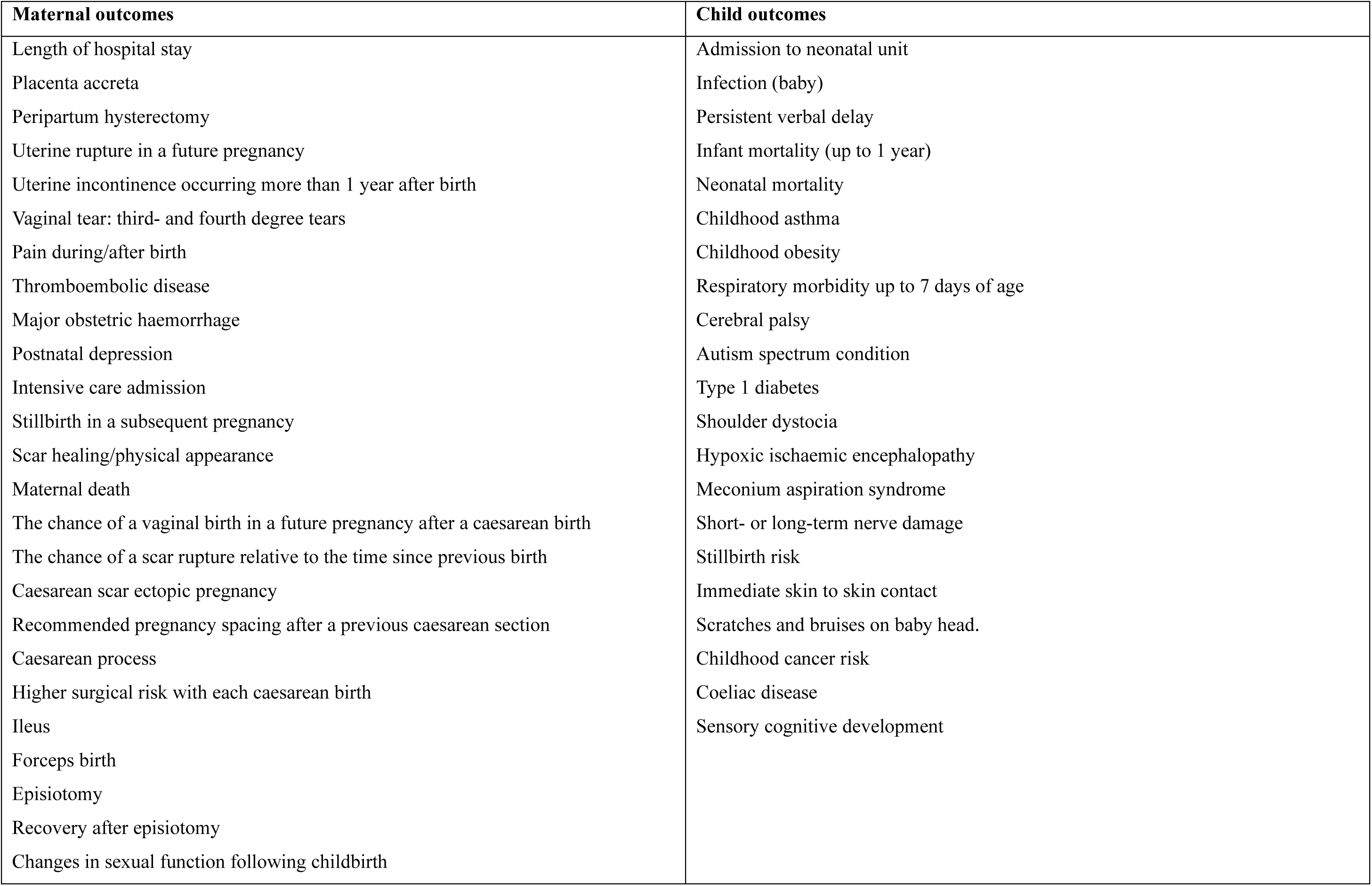
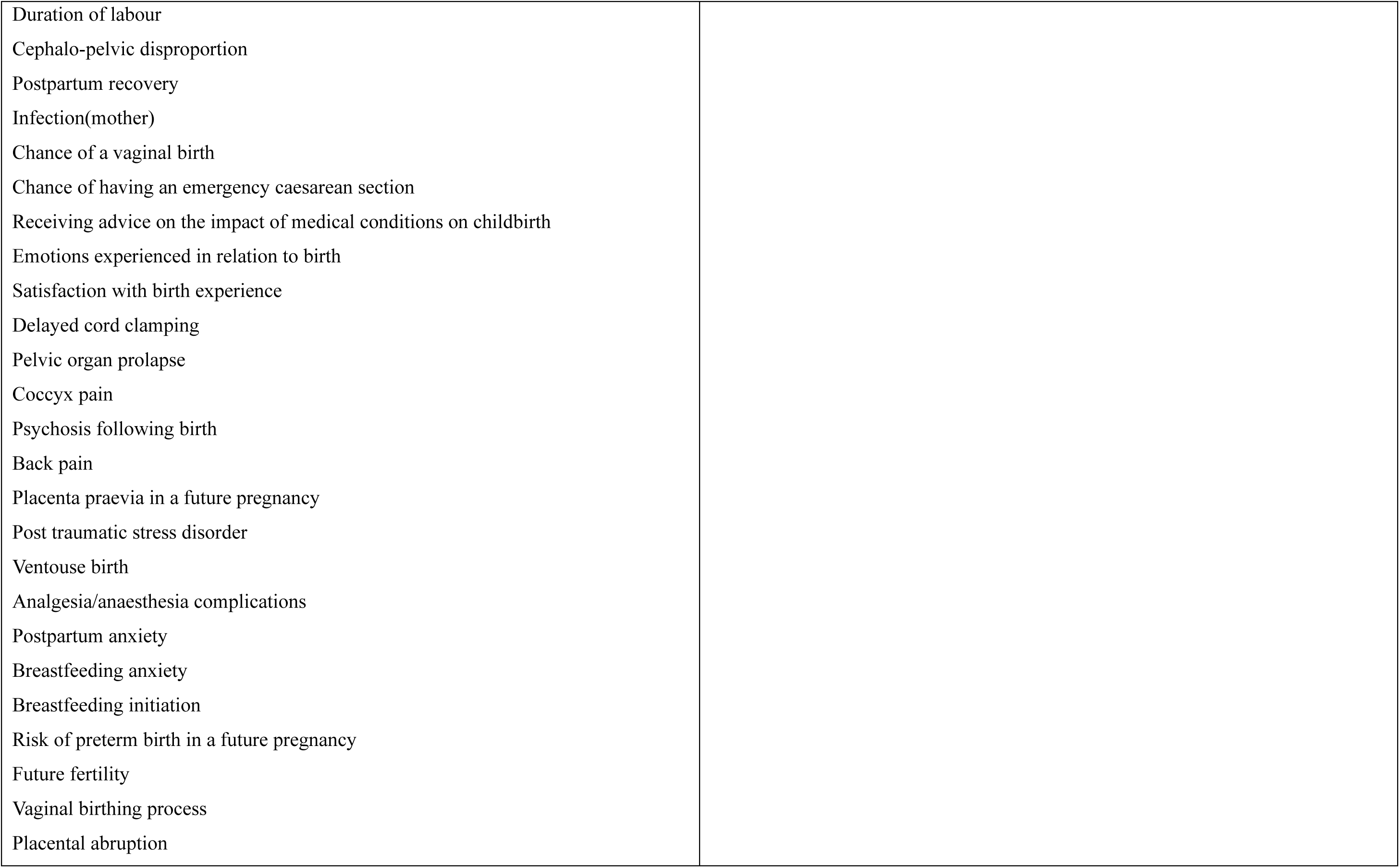
All outcomes as presented in the Delphi study.

## Notes

### Author Declarations

Ethics committee of East of Scotland Research Ethics Service and the Health Research Authority (HRA) and Health and Care Research Wales (HCRW) gave ethical approval for this work.

